# Assessing the health risk of living near composting facilities on lung health, fungal and bacterial disease in cystic fibrosis: a UK CF Registry study

**DOI:** 10.1101/2022.05.08.22274725

**Authors:** Muhammad Saleem Khan, Philippa Douglas, Anna L. Hansell, Nicholas J. Simmonds, Frédéric B. Piel

## Abstract

**Aim:** To explore the health risk of living near permitted composting sites (PCSs) on disease severity in children and adults with cystic fibrosis (CF) across the UK.

**Methods:** A semi-individual cross-sectional study was used to examine risk of disease severity in people with CF (pwCF) within and beyond 4 km of PCSs in the UK in 2016. All pwCF registered in the UK CF Registry were eligible for this study. Linear and Poisson regressions, adjusted for age, gender, genotype, BMI, *Pseudomonas aeruginosa (P.aeruginosa)* and deprivation, were used to quantify associations between distance to a PCS and percent predicted forced expiratory volume in one second (ppFEV_1_), pulmonary exacerbations (#IVdays), and fungal and bacterial infections.

**Results:** The mean age of the 9,361 pwCF (3,931 children and 5,430 adults) studied was 20.1 (*SD*=14.1) years; 53.3% were male; and 49.2% were homozygous *F508del*. Over 10% of pwCF (n=1,015) lived within 4 km of a PCS. We found no statistically significant difference in ppFEV_1_ and #IVdays/year in children. However, in adults, ppFEV_1_ was −1.07% lower (95% confidence interval (CI): −2.29%, 0.16 %) and #IVdays/year were 1.02 day higher (95%CI: 1.01, 1.04) within 4 km of a PCS. Furthermore, there were statistically significant differences in mean ppFEV_1_ in CF adults with *Aspergillus fumigatus* (54.5% *vs* 59.7%, p=0.001) *and Candida spp*. (54.8% *vs* 59.0%, p=0.001) residing within 4 km of a PCS. No associations were identified for allergic bronchopulmonary aspergillosis, *P.aeruginosa* or *Staphylococcus aureus*.

**Conclusions:** This novel national study provides evidence that adults with CF living near a PCS may experience small reductions in lung function, an increased risk of pulmonary exacerbations, and more frequent fungal infections. If confirmed by studies using refined exposure assessment methods accounting for bioaerosol dispersion, these results could have important implications for the living environment of pwCF.

## Background

Cystic fibrosis (CF) is an autosomal recessive disorder caused by mutations in the CF transmembrane conductance regulator (CFTR) gene which is located on the long arm of chromosome 7. It is one of the most commonly inherited life-shortening diseases in Caucasians with a birth prevalence of one in every 2,500 live births in the UK (UK CF Registry., 2020). Almost 500 variants in *CFTR* have been identified so far (CFTR2 database, 2021). *CFTR* protein functions predominantly as a chloride channel in epithelial cells, regulating the movement of salt and water in and out of cells, resulting in clinical complications particularly affecting the lungs and the gastrointestinal system. People with CF (pwCF) produce thick secretions in the lung which provide a suitable environment for the growth of pathogens, including bacteria and fungi. This leads to recurrent respiratory infections, commonly termed pulmonary exacerbations (PEx), and an inflammatory environment resulting in progressive respiratory disease (Ranganathan et al., 2013).

Respiratory infections are one of the leading causes of morbidity and mortality among the CF population. Substantial work has been done on the treatment and prevention of bacterial infection in pwCF (Chiappini et al., 2014; Gilligan, 2014). This has contributed to measurable increases in life expectancy but also may result in an increased risk for colonisation of fungi (Gilligan, 2014). Fungi are ubiquitous organisms and the most common fungus among pwCF is *Aspergillus fumigatus (A.fumigatus)* (Burgel et al., 2016b). Based on data from the UK CF Registry, the prevalence of positive cultures for *A.fumigatus* increased from 6% in 2007 to 15% in 2016 (UK CF Registry., 2020). Evidence suggests that some fungal infections are associated with a greater decline in lung function and an increased rate of PEx (Saunders et al., 2016). The mechanisms underlying this trend remain unclear, but a substantial contribution from environmental factors seems likely (Crawford et al., 2015; Mendell et al., 2011).

A better understanding of the risk of infections around composting facilities might help to better identify important environmental risk factors and their impact on the health of pwCF (Robertson et al., 2019). There are some public health concerns related to waste composting as it results in airborne bioaerosols which are released during the composting process (Douwes et al., 2003). The number of permitted composting sites (PCSs) more than doubled between 2010 and 2017 in England (Douglas et al., 2021). Concentrations of bioaerosols are higher at PCSs when the compost is agitated (e.g. shredded, turned and screened) (Taha et al., 2006). These bioaerosols include fungi and fungal spores, gram-negative and spore-producing gram-positive bacteria (e.g., actinomycetes), endotoxins and other various particulates sized between 0.2 to 100 µm in diameter (Douwes et al., 2003). The majority of bioaerosols emitted from composting facilities are small (<3 µm in diameter) and therefore can be inhaled and penetrate the lungs (Robertson et al., 2019). Although previous studies have shown that bioaerosol concentrations fall within 250m below levels considered acceptable by the UK Environment Agency’s risk assessment (Environment Agency and Department for Environment, 2016), there is evidence that some bioaerosols may not ground over distances of a few kilometres from a PCS (Williams et al., 2019a). Furthermore, the role of these bioaerosols on the health of vulnerable population sub-groups remains unclear. It should be noted that, at present, there are no quantitative dose-response estimates to inform legislation on bioaerosol emissions from PCSs.

Occupational studies have shown that exposure to fungi, bacteria and particulates, especially those <10 µm diameter, affects human health (Mack et al., 2019). These bioaerosols can penetrate the alveolar sacs of the lung, causing damage and loss of lung function, resulting in respiratory complications. These complications include allergic asthma, rhinitis, hypersensitivity pneumonitis, allergic bronchopulmonary aspergillosis (ABPA), eye and skin irritations, bronchitis, airway obstruction such as chronic obstructive pulmonary disease (COPD), organic dust toxic syndrome and toxic pneumonitis. The evidence of the possible associations between bioaerosol emissions from composting sites and health effects in the surrounding community is limited, although potential risks cannot be completely ruled out (Douglas et al., 2016; Robertson et al., 2019; Roca-Barcelo et al., 2020).

People with lung diseases such as COPD, asthma and CF are likely to be more vulnerable to the harmful effects of living near a PCS, and exposure to bioaerosols may represent a particular health risk among pwCF (Walser et al., 2015). This national-scale study aimed to investigate whether proximity to PCS - as a bioaerosol exposure proxy - is associated with a lower lung function, higher number of PEx, and more frequent fungal and bacterial infections in pwCF. This is based on the hypothesis that bioaerosols emitted from PCSs may be a source of such pathogens, including *A.fumigatus*.

## Methods

### Study design and population

We used a semi-individual cross-sectional study design to investigate the impact of living near a PCS on lung function, PEx, and fungal and bacterial disease among children and adults with CF. Our health outcome data came from the UK CF Registry; a secure database that collects annual data on approximately 10,000 patients from all CF specialist centres across the UK, representing >99% of pwCF (Taylor-Robinson et al., 2018). Registry records are based on annual assessments of each individual, including data on demography, health outcomes and lifestyle factors, collected at CF centres across the UK, which are then centralised into one big database at the UK CF Registry. The present study focuses on all pwCF registered in the UK CF Registry who had an annual review conducted in 2016. In the UK, pwCF transition from paediatric to adult care occurs at around the age of 16. Individuals aged less than 16 years (0-15) were therefore considered as children, and those aged 16 and above as adults. Most analyses were conducted separately for children and adults. Each individual was located based on their full residential postcode (postcode unit). Postcodes are regularly checked and updated in the UK CF Registry. In England and Wales, a postcode unit has an average of 43 (SD=38, median=33) residents and 18 (SD=15, median=14) occupied households (ONS, 2013). Coordinates of postcode units were obtained from the Ordnance Survey’ Code-Point Open dataset (Ordnance Survey., 2022).

### Outcome variables

We considered three key outcome variables for this analysis: i) lung function, ii) PEx and iii) evidence of fungal and bacterial disease. Lung function was measured as the percent predicted forced expired volume in one second (ppFEV_1_) as this is a key prognostic measure in CF (Szczesniak et al., 2017). ppFEV_1_ was calculated using the Global Lung Function Initiative predictive equation (Quanjer et al., 2012). Children under the age of six were included in the PEx analysis, but not in the lung function analysis (see below). Figure 1 provides a schematic overview of our inclusion and exclusion criteria.

**Figure 1:**
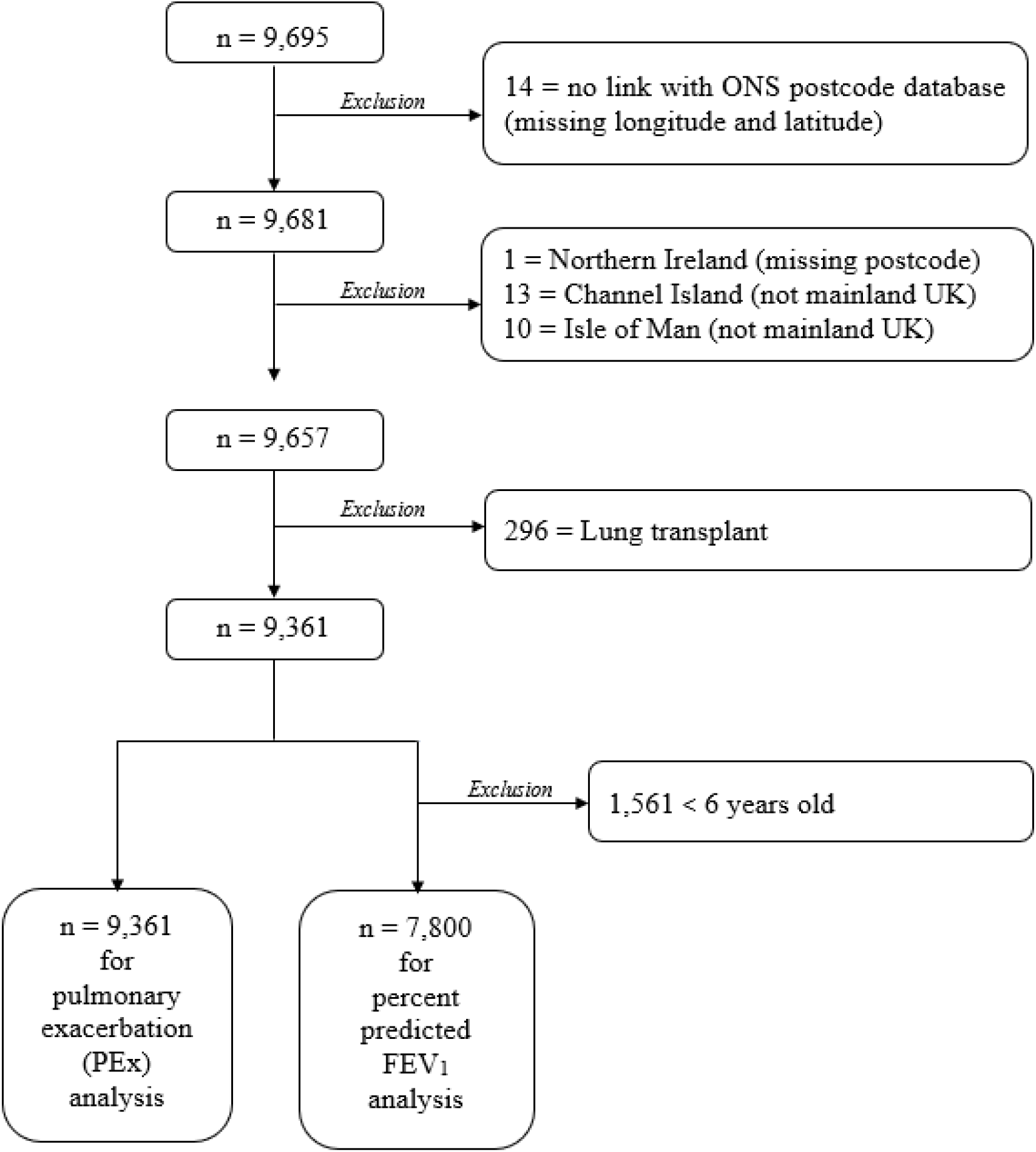
Flow diagram of the inclusion and exclusion criteria used for people with CF (pwCF) in the 2016 UK CF Registry database.

There is no standard definition of a PEx, however, the use of intravenous (IV) antibiotics (which are prescribed to treat a PEx) is widely considered to be a good proxy in observational studies. Here, a PEx was defined as the use of IV antibiotics at home or in hospital. We used the total number of days on IV antibiotics (home and hospital IV courses) received for a Pex during the twelve months preceding the annual review as we considered it to be a more robust and reliable measure of disease severity than the number of antibiotic courses (Hoo et al., 2018). Spirometry is not reliable under six years of age (i.e. < 6 years old) due to the inability of the child to perform the standard pulmonary function tests accurately (Kerem et al., 2014). Therefore, for our analysis of ppFEV_1_, all pwCF younger than six years old (n=1,561) were excluded. Individuals who had a lung transplant before 2017 (n=296) were also excluded as their transplanted lungs and immunocompromised state would confound results.

To define the presence of fungal infection or disease, we used records of positive fungal cultures (sputum, cough/throat swab or bronchoalveolar lavage) of *A.fumigatus* and *Candida spp*. in the year preceding the annual review and/or a record of ABPA. In the UK CF Registry, ABPA is defined using standard international diagnostic criteria (see Appendix A) (Stevens et al., 2003). We also considered two common bacterial infections recorded in the UK CF Registry: *Pseudomonas aeruginosa (P.aeruginosa)* and *Staphylococcus aureus (S.aureus)* using positive record of culture in the year preceding the annual review.

### Exposure variable

In the UK, commercial-scale composting facilities need a permit to operate. In England, such a permit is required if the facility is dealing with over 60 tonnes of compost at any one time (DEFRA, 2014), but different tonnages are used in Scotland, Wales and Northern Ireland. All operational PCSs in the UK at the end of 2016 were identified using information provided by the UK Environment Agency (England), Natural Resources Wales, the Scottish Environment Protection Agency and the Northern Ireland Environment Agency. The sites were geocoded in a Geographic Information System (ArcGIS 10.7.1, ESRI, Redlands, CA) using the postcode of the facilities provided by the Environment Agencies. Figure 2 illustrates the distribution of PCSs across the UK. Distance between the nearest PCS and the pwCF’s postcode was measured using Euclidian distances (point distance analysis) calculated in ArcGIS. In line with previous studies (Douglas et al., 2016; Robertson et al., 2019; Roca-Barcelo et al., 2020), we considered a range of circular distance bands (0 – 250 m, >250 - ≤750 m, >750 - ≤1.5 km, >1.5 - ≤ 2.5 km, >2.5 - ≤ 4.0 km and > 4.0 km) around each PCS (Table S1 and Appendix B). Ultimately, because the small number of pwCF living near PCS limited the statistical power of analyses in smaller bands, we focussed our study on within and beyond 4 km.

**Figure 2:**
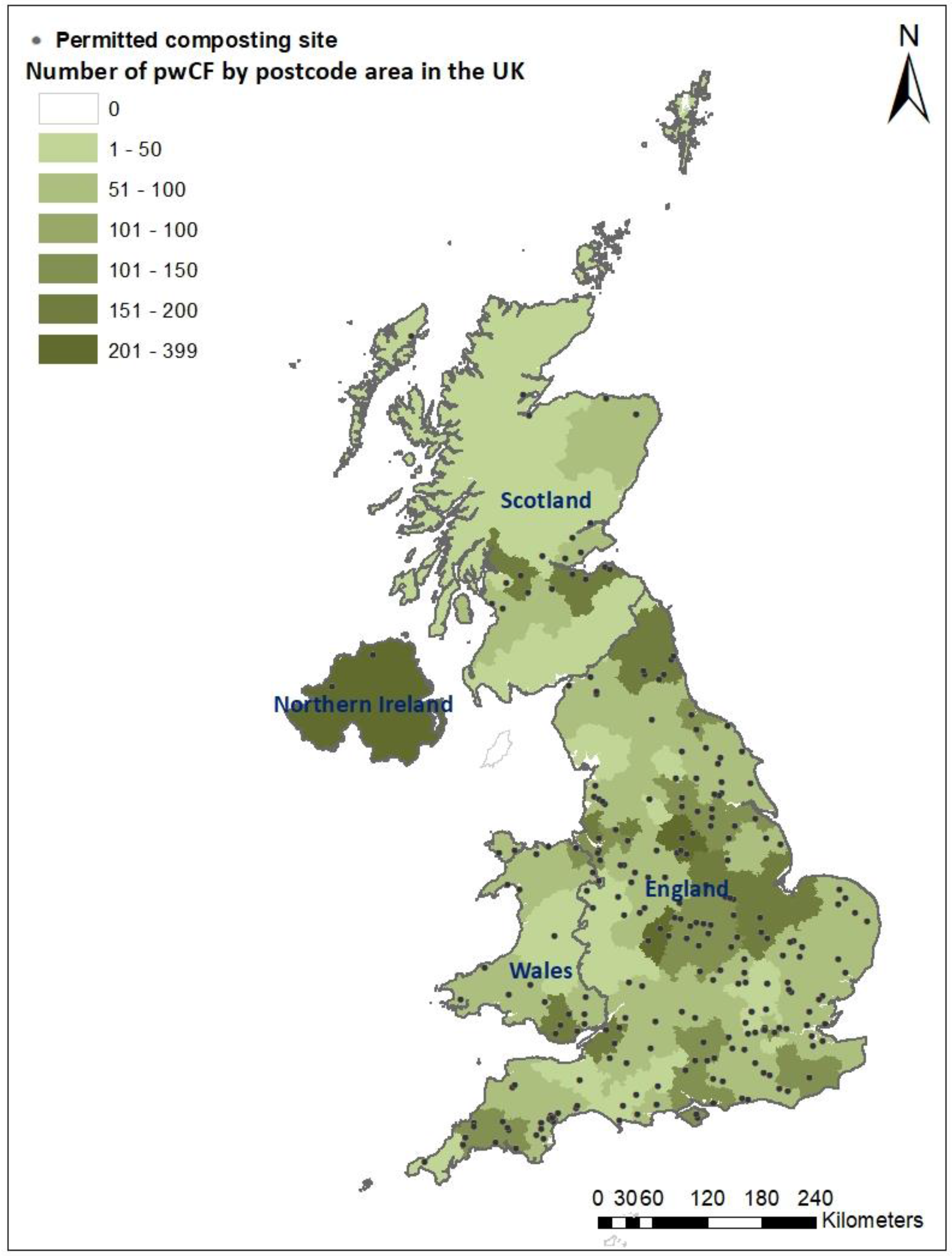
Map of the number of pwCF by postcode area and permitted composting sites (PCSs), in the UK in 2016 based on the centroid of postcodes provided by the Environment Agencies of England (n=209), Scotland (n=21), Wales (n=24) and Northern Ireland (n=02).

### Covariates

We chose *a priori* the following variables as potential confounders: age, sex, body mass index (BMI), *CFTR* genotype, *P.aeruginosa*, urban-rural classification and socio-economic status (SES). We categorised the age variable into two groups: children (<16 years) and adults (≥16 years) (Lalley, 2013; Vaz Fragoso et al., 2016). We used a binary variable for gender (M *vs* F) in our analysis. BMI is an important parameter to assess the nutritional status of CF individuals. We used absolute BMI in adults and BMI percentile, calculated based on a age, sex, height and weight, in children (Vogel M., 2022). Maintaining a BMI above the 50^th^ percentile for children and an absolute BMI >19 kg/m^2^ in adults is associated with better outcomes in CF (Stenbit and Flume, 2011). The Office for National Statistics (ONS) postcode classification was used to assign an urban or rural designation to each pwCF residential postcode (see Appendix C) (ONS, 2020). We used the 2011 UK Townsend deprivation score quintiles at output area level (highest resolution available) as a proxy of the SES of pwCF (ONS, 2020; Taylor-Robinson et al., 2013); the first quintile (Q1) referring to the least deprived areas of the UK whereas the fifth quintile (Q5) corresponds to the most deprived. Infections with *P.aeruginosa* was defined by a positive culture in the year preceding the annual review.

### *CFTR* genotype

Two ways of classifying *CFTR* mutations have commonly been used in the past. First, six (or seven) classes based on protein synthesis and function (De Boeck and Amaral, 2016). This classification system has significant limitations as there is considerable overlap between the classes and not all mutations have been studied sufficiently to be able to classify them. Second, a simplified classification using the common *F508del* mutation status as the main discriminator – i.e. homozygous *F508del*, heterozygous *F508del* and ‘other.’ This classification does not fully take into account the clinical phenotype as the ‘other’ category covers a wide spectrum of disease severity. To improve on these classifications and make it more relevant to contemporary practice, we devised a new eight group classification (Table S2) based on the different combinations of the recognised terms developed in parallel with new *CFTR*-targeted therapies – so-called *CFTR*-modulators: residual function (RF), minimal function (MF) and gating function (Heijerman et al., 2019; Middleton et al., 2019). We believe that an added benefit of this new classification is that it aligns better with potential treatment options for patients.

### Statistical analysis

Spatial and statistical analyses were performed using ArcGIS 10.7.1 (ESRI, Redlands, CA) and *R* 4.1.1 (Team, 2017), respectively. Individual characteristics including age, sex, *CFTR* genotype class and *P.aeruginosa* status, and area-level descriptors such as deprivation, rural-urban classification and country were compared among pwCF residing within and beyond 4 km from a PCS using two-way ANOVAs. Separate regression models were built for children and adults. A multiple linear regression analysis was run to explore the association between ppFEV_1_ and distance (≤ 4 km *vs* > 4 km) from a PCS. Poisson regression was employed to explore the association between the total #IVdays for PEx and distance from a PCS. The models were adjusted in a forward-stepwise approach with covariates added one-by-one using the Akaike information criterion (AIC) (Burnham and Anderson, 2003). The best model was identified based on the lowest AIC (Table S3). A p-value of 0.05 was considered statistically significant and 95% corresponding confidence intervals (CIs) and p-values are reported. Fungal and bacterial infection rates for *A.fumigatus, Candida spp*., ABPA, *P.aeruginosa and S.aureus* were calculated among pwCF within and beyond 4km of a PCS. An ANCOVA was used to calculate adjusted mean ppFEV_1_, separately in children and adults, for each of these five measurements of fungal and bacterial disease.

### Ethics and approval

This study used individual-level data pseudonymised by the UK CF Registry. The study was approved by the UK CF Trust and by SAHSU’s Liaison Committee and PHE’s Environmental Public Health Programme Board. Data were stored and processed on a secure ISO 27001:2013 server.

## Results

A total of 9,695 pwCF had an annual review in 2016. Data for 9,631 individuals, including 3,931 children and 5,430 adults, were available for our PEx and fungal and bacterial disease analysis. After removal of children <6 years old, 2,370 children remained for our ppFEV1 analysis. Table 1 compares the demographic characteristics of pwCF residing within and beyond 4 km from a PCS. The average age of pwCF was 20.1 (*SD*=14.1) years; 49% (n=4,588) were aged 16-40 years; 53.3% were male and 19.2% lived in rural areas. There was an almost equal proportion of pwCF across all quintiles of deprivation. The geographical distribution of pwCF by country was as follows: England = 7,675 (82.0%), Wales = 494 (5.3%), Scotland = 796 (8.5%) and Northern Ireland = 396 (4.2%) (Figure 2). In terms of *CFTR* genotypes, 49.2% were homozygous for F508del, 24.3% had at least one MF mutation, 11.4% had a RF mutation and 6.2% had a gating mutation (Table S2). In 2016, there were 256 active PCSs in the UK (209 in England; 24 in Wales; 21 in Scotland; and 2 in Northern Ireland) (Figure 2); 1,015 pwCF (10.8%) lived within 4 km of a PCS.

**Table 1:**
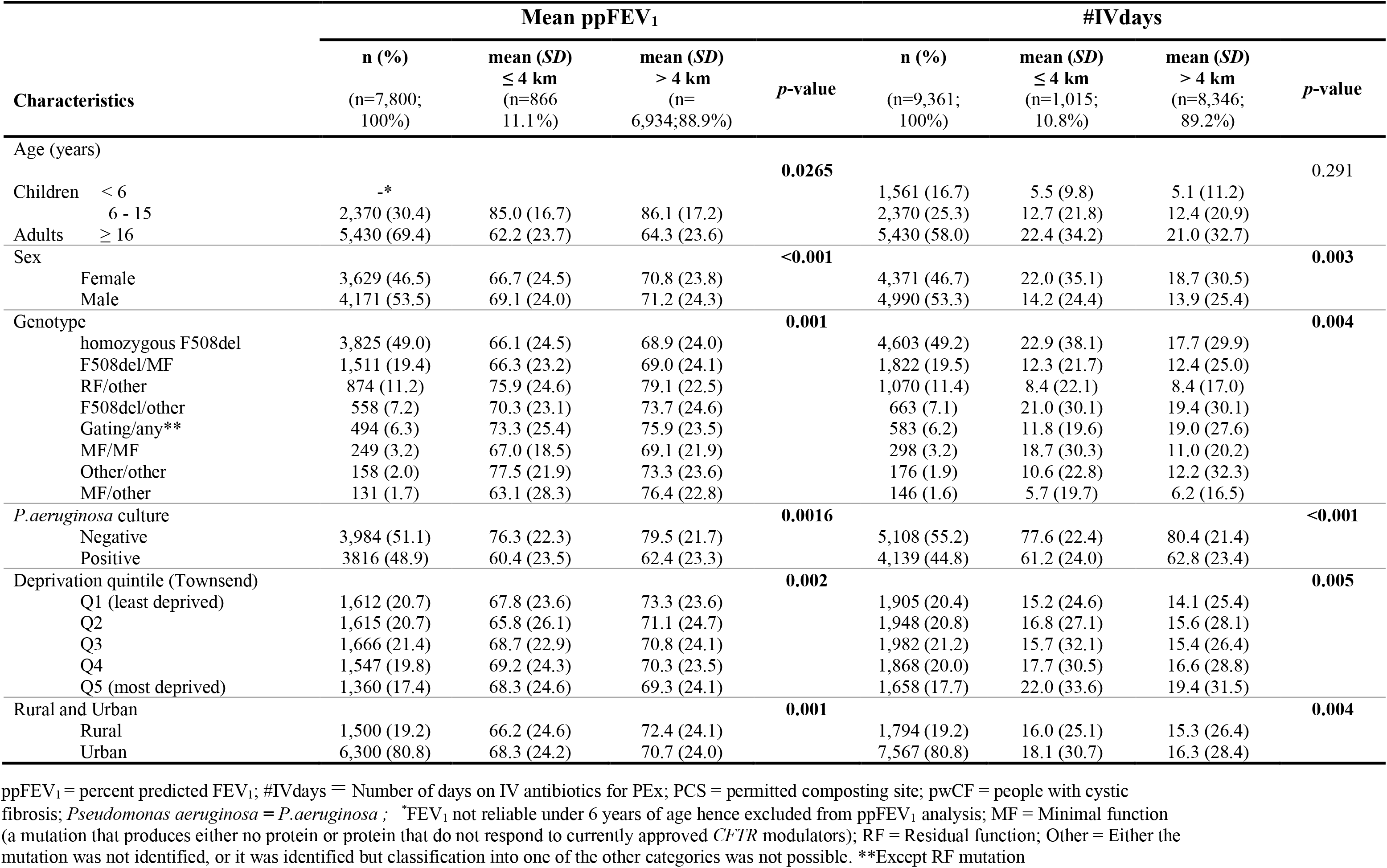
Mean ppFEV_1_ and #IVdays within and beyond 4 km of a PCS in pwCF in the UK in 2016. p-value <0.05 are shown in bold.

| Characteristics | Mean ppFEV<sub>1</sub> |  |  | p-value | #IVdays |  |  | p-value |
| --- | --- | --- | --- | --- | --- | --- | --- | --- |
|  | n (%) | mean (SD)<br>≤ 4 km | mean (SD)<br>> 4 km |  | n (%) | mean (SD)<br>≤ 4 km | mean (SD)<br>> 4 km |  |
|  | (n=7,800;<br>100%) | (n=866<br>11.1%) | (n=6,934;88.9%) |  | (n=9,361;<br>100%) | (n=1,015;<br>10.8%) | (n=8,346;<br>89.2%) |  |
| Age (years) |  |  |  | **0.0265** |  |  |  | 0.291 |
| Children < 6 | _* |  |  |  | 1,561 (16.7) | 5.5 (9.8) | 5.1 (11.2) |  |
| 6 - 15 | 2,370 (30.4) | 85.0 (16.7) | 86.1 (17.2) |  | 2,370 (25.3) | 12.7 (21.8) | 12.4 (20.9) |  |
| Adults ≥ 16 | 5,430 (69.4) | 62.2 (23.7) | 64.3 (23.6) |  | 5,430 (58.0) | 22.4 (34.2) | 21.0 (32.7) |  |
| Sex |  |  |  | **<0.001** |  |  |  | **0.003** |
| Female | 3,629 (46.5) | 66.7 (24.5) | 70.8 (23.8) |  | 4,371 (46.7) | 22.0 (35.1) | 18.7 (30.5) |  |
| Male | 4,171 (53.5) | 69.1 (24.0) | 71.2 (24.3) |  | 4,990 (53.3) | 14.2 (24.4) | 13.9 (25.4) |  |
| Genotype |  |  |  | **0.001** |  |  |  | **0.004** |
| homozygous F508del | 3,825 (49.0) | 66.1 (24.5) | 68.9 (24.0) |  | 4,603 (49.2) | 22.9 (38.1) | 17.7 (29.9) |  |
| F508del/MF | 1,511 (19.4) | 66.3 (23.2) | 69.0 (24.1) |  | 1,822 (19.5) | 12.3 (21.7) | 12.4 (25.0) |  |
| RF/other | 874 (11.2) | 75.9 (24.6) | 79.1 (22.5) |  | 1,070 (11.4) | 8.4 (22.1) | 8.4 (17.0) |  |
| F508del/other | 558 (7.2) | 70.3 (23.1) | 73.7 (24.6) |  | 663 (7.1) | 21.0 (30.1) | 19.4 (30.1) |  |
| Gating/any** | 494 (6.3) | 73.3 (25.4) | 75.9 (23.5) |  | 583 (6.2) | 11.8 (19.6) | 19.0 (27.6) |  |
| MF/MF | 249 (3.2) | 67.0 (18.5) | 69.1 (21.9) |  | 298 (3.2) | 18.7 (30.3) | 11.0 (20.2) |  |
| Other/other | 158 (2.0) | 77.5 (21.9) | 73.3 (23.6) |  | 176 (1.9) | 10.6 (22.8) | 12.2 (32.3) |  |
| MF/other | 131 (1.7) | 63.1 (28.3) | 76.4 (22.8) |  | 146 (1.6) | 5.7 (19.7) | 6.2 (16.5) |  |
| *P.aeruginosa* culture |  |  |  | **0.0016** |  |  |  | **<0.001** |
| Negative | 3,984 (51.1) | 76.3 (22.3) | 79.5 (21.7) |  | 5,108 (55.2) | 77.6 (22.4) | 80.4 (21.4) |  |
| Positive | 3816 (48.9) | 60.4 (23.5) | 62.4 (23.3) |  | 4,139 (44.8) | 61.2 (24.0) | 62.8 (23.4) |  |
| Deprivation quintile (Townsend) |  |  |  | **0.002** |  |  |  | **0.005** |
| Q1 (least deprived) | 1,612 (20.7) | 67.8 (23.6) | 73.3 (23.6) |  | 1,905 (20.4) | 15.2 (24.6) | 14.1 (25.4) |  |
| Q2 | 1,615 (20.7) | 65.8 (26.1) | 71.1 (24.7) |  | 1,948 (20.8) | 16.8 (27.1) | 15.6 (28.1) |  |
| Q3 | 1,666 (21.4) | 68.7 (22.9) | 70.8 (24.1) |  | 1,982 (21.2) | 15.7 (32.1) | 15.4 (26.4) |  |
| Q4 | 1,547 (19.8) | 69.2 (24.3) | 70.3 (23.5) |  | 1,868 (20.0) | 17.7 (30.5) | 16.6 (28.8) |  |
| Q5 (most deprived) | 1,360 (17.4) | 68.3 (24.6) | 69.3 (24.1) |  | 1,658 (17.7) | 22.0 (33.6) | 19.4 (31.5) |  |
| Rural and Urban |  |  |  | **0.001** |  |  |  | **0.004** |
| Rural | 1,500 (19.2) | 66.2 (24.6) | 72.4 (24.1) |  | 1,794 (19.2) | 16.0 (25.1) | 15.3 (26.4) |  |
| Urban | 6,300 (80.8) | 68.3 (24.2) | 70.7 (24.0) |  | 7,567 (80.8) | 18.1 (30.7) | 16.3 (28.4) |  |

ppFEV<sub>1</sub> = percent predicted FEV<sub>1</sub>; #IVdays = Number of days on IV antibiotics for PEx; PCS = permitted composting site; pwCF = people with cystic fibrosis; *Pseudomonas aeruginosa* = *P.aeruginosa*; \*FEV<sub>1</sub> not reliable under 6 years of age hence excluded from ppFEV<sub>1</sub> analysis; MF = Minimal function (a mutation that produces either no protein or protein that do not respond to currently approved *CFTR* modulators); RF = Residual function; Other = Either the mutation was not identified, or it was identified but classification into one of the other categories was not possible. \*\*Except RF mutation

### Lung function

Overall, the mean ppFEV_1_ was 85.0 within 4 km of a PCS *vs* 86.1 beyond 4 km (p=0.391) and 62.2 within 4 km *vs* 64.3 beyond 4km (p=0.044) in children and adults, respectively. The age (p= 0.021), sex (p <0.001), deprivation index (p= 0.002), rural proportion (p= 0.001) and *CFTR* genotype (p= 0.001) characteristics of pwCF residing within 4 km from a PCS were all statistically significantly different to those living farther away from such facilities (Table 1). There was no statistically significant difference in mean ppFEV1 between children 6-15 years old with CF who lived within and beyond 4 km of a PCS (Table 2). In our unadjusted linear regression analysis, we found a −1.44% (95% CI: −2.84, −0.04) lower ppFEV_1_ in adults with CF residing within 4 km of a PCS compared to the rest of pwCF. This difference in ppFEV_1_ became smaller and the association was attenuated −1.07% (95% CI: −2.29, −0.16, p=0.088) after adjusting for age, sex, BMI, genotype, *P.aeruginosa*, and deprivation (Table 2).

**Table 2:** Linear and Poisson regression analysis of the relationship between ppFEV_1_ and #IVdays to residential postcode distance from PCS among children and adults with CF in the UK in 2016. p-value <0.05 are shown in bold.

| Residential<br>postcode<br>distance | ppFEV<sub>1</sub> |  |  |  |  | #IVdays |  |  |  |  |  |
| --- | --- | --- | --- | --- | --- | --- | --- | --- | --- | --- | --- |
|  | mean<br>ppFEV<sub>1</sub> | Coeff. | 95% CI | *p*-<br>*value* | AIC | mean<br>#IVdays | Coeff. | Exp (B) | 95% CI | *p*-<br>*value* | AIC |
| **Children (6 -15 years) (n=2,370)** |  |  |  |  |  | **Children (0 -15 years) (n=3,931)** |  |  |  |  |  |
| **Unadjusted** |  |  |  | 0.391 | 19297.9 |  |  |  |  | 0.087 | 103952.3 |
| ≤ 4 km | 85.0 | -0.75 | -2.46, 0.96 |  |  | 9.8 | 0.02 | 1.02 | 0.99, 1.05 |  |  |
| > 4 km | 86.1 | 1.00 | - |  |  | 9.5 |  | 1.00 | - |  |  |
| **Adjusted\*** |  |  |  | 0.358 | 18887.6 |  |  |  |  | 0.623 | 84834.0 |
| ≤ 4 km | 86.0** | -0.74 | -2.31, 0.83 |  |  | 10.3 | 0.01 | 1.01** | 0.99, 1.04 |  |  |
| > 4 km | 86.8** | 1.00 | - |  |  | 10.3 |  | 1.00** |  |  |  |
| **Adults (16 - 86 years) (n=5,430)** |  |  |  |  |  | **Adults (16 - 86 years) (n=5,430)** |  |  |  |  |  |
| **Unadjusted** |  |  |  | **0.044** | 47438.5 |  |  |  |  | **<0.001** | 231519.0 |
| ≤ 4 km | 62.2 | -1.44 | -2.84, -0.04 |  |  | 22.4 | 0.04 | 1.04 | 1.03, 1.06 |  |  |
| > 4 km | 64.3 | 1.00 | - |  |  | 21.0 |  | 1.00 | - |  |  |
| **Adjusted\*** |  |  |  | 0.088 | 46040.1 |  |  |  |  | **<0.001** | 187492.7 |
| ≤ 4 km | 64.4** | -1.07 | -2.29, 0.16 |  |  | 19.4 | 0.02 | 1.02** | 1.01, 1.04 |  |  |
| > 4 km | 66.3** | 1.00 | - |  |  | 18.4 |  | 1.00** |  |  |  |

ppFEV<sub>1</sub> = percent predicted FEV<sub>1</sub>; #IVdays = Number of days on IV antibiotics for PEx; pwCF = people with cystic fibrosis; PCS = permitted composting site
\*Adjusted for age, sex, BMI (children model adjusted for BMI percentile and adult model adjusted for absolute BMI), genotype, *P.aeruginosa* and deprivation
\*\* calculated using ANCOVA (Analysis of Covariance) adjusting for age, sex, BMI (children model adjusted for BMI percentile and adult model adjusted for absolute BMI), genotype, *P.aeruginosa* and deprivation

### Pulmonary exacerbations

The mean number of #IVdays were 9.8 *vs* 9.5 days per year in children living within and beyond 4km from a PCS whereas in adult the mean number of #IVdays were 22.4 *vs* 21.0 days per year. In univariate analyses, there was a statistically significant difference in #IVdays per year between males and females (p= <0.003), deprivation quintiles (p= 0.005), rural and urban areas (p= 0.004), genotype category (p= 0.004) for pwCF residing within and beyond 4 km of the PCS (Table 1). We found no statistically significant difference in #IV days between children with CF who lived within 4 km of a PCS and those who lived farther away. Using Poisson regression analysis in adults, we identified an increase of 1.04 (95%CI: 1.03-1.06) and 1.02 (95%CI: 1.01,1.04) in the rate of #IVdays per year before and after adjustment for age, sex, BMI, genotype, *P.aeruginosa* and deprivation (Table 2).

### Fungal and bacterial infections

Table 3 shows that there was no difference in the point prevalence of *A.fumigatus* positive cultures within and beyond 4km of a PCS when considering all pwCF (15.3% *vs* 15.5%, respectively). However, there was a statistically significant difference in the mean ppFEV_1_ in adults with CF with *A.fumigatus* positive culture within and beyond 4km of PCS (55.9% *vs* 62.1%: p=0.001). This difference remained significant after adjusting for age, sex, BMI, genotype category and deprivation (54.5% *vs* 59.7%; p=0.001). Furthermore, the point prevalence of *Candida spp*. was 3.6% higher (p=0.01) close to PCSs. The difference of mean ppFEV_1_ increased to 4.2% (54.8% *vs* 59.0%; p=0.042) in the adjusted multivariate analysis for adults. No statistically significant differences were found in the point prevalence of ABPA, *P.aeruginosa* or *S.aureus* within and beyond 4 km from a PCS in adults with CF. In addition, there was no statistically significant difference in the mean ppFEV_1_ of children or adults with CF who live within and beyond 4 km of a PCS in the UK when considering cultures for ABPA, *P.aeruginosa* or *S.aureus* (Table 3).

**Table 3:** Respiratory microbiology, fungal disease and mean ppFEV_1_ among children and adults with CF within and beyond 4km of PCS in the UK in 2016. p-value <0.05 are shown in bold.

|  | Distance band<br>Crude |  |  | Distance band<br>Adjusted* |  |  |
| --- | --- | --- | --- | --- | --- | --- |
|  | ≤ 4 km | > 4 km | p-value | ≤ 4 km | > 4 km | p-value |
| Total Number of pwCF (n) | 866 | 6,934 | - | - | - | - |
| Number of pwCF who had culture*<br>(n) | 842 | 6,730 | - | - | - | - |
| *A.fumigatus* |  |  |  |  |  |  |
| *n* (point prevalence %) | 129 (15.3) | 1,041 (15.5) | 0.911 |  |  |  |
| mean ppFEV<sub>1</sub> |  |  |  |  |  |  |
| Children | 77.2 | 79.0 | 0.662 | 78.3 | 80.4 | 0.631 |
| Adults | 55.9 | 62.1 | **0.001** | 58.2 | 62.0 | **0.005** |
| ABPA |  |  |  |  |  |  |
| *n* (point prevalence %) | 76 (8.8) | 551 (7.9) | 0.397 |  |  |  |
| mean ppFEV<sub>1</sub> |  |  |  |  |  |  |
| Children | 81.2 | 75.3 | 0.229 | 86.6 | 82.6 | 0.179 |
| Adults | 60.2 | 59.1 | 0.717 | 60.3 | 60.1 | 0.696 |
| *Candida spp.* |  |  |  |  |  |  |
| *n* (point prevalence %) | 180 (21.4) | 1,195 (17.8) | **0.010** |  |  |  |
| mean ppFEV<sub>1</sub> |  |  |  |  |  |  |
| Children | 79.6 | 79.3 | 0.929 | 84.0 | 82.9 | 0.921 |
| Adults | 53.6 | 57.8 | **0.048** | 56.9 | 59.9 | **0.029** |
| *P.aeruginosa* |  |  |  |  |  |  |
| *n* (point prevalence %) | 439 (52.1) | 3,377 (50.2) | 0.284 |  |  |  |
| mean ppFEV<sub>1</sub> |  |  |  |  |  |  |
| Children | 80.5 | 82.3 | 0.436 | 80.9 | 84.1 | 0.399 |
| Adults | 56.8 | 58.1 | 0.280 | 58.5 | 59.6 | 0.240 |
| *S.aureus* |  |  |  |  |  |  |
| *n* (point prevalence %) | 76 (8.3) | 558 (9.0) | 0.469 |  |  |  |
| mean ppFEV<sub>1</sub> |  |  |  |  |  |  |
| Children | 71.2 | 76.1 | 0.541 | 63.7 | 70.8 | 0.472 |
| Adults | 58.0 | 56.9 | 0.690 | 59.1 | 59.0 | 0.663 |

*Aspergillus fumigatus* = *A.fumigatus* ; *Pseudomonas aeruginosa* = *P.aeruginosa* ; *Staphylococcus aureus* = *S.aureus*
\*(Sputum/ Cough Swab/ Bronchoscopy); ppFEV<sub>1</sub> = percent predicted with FEV<sub>1</sub>; pwCF = people with cystic fibrosis; ABPA = Allergic Bronchopulmonary Aspergillosis; PCS = permitted composting site; Children = 6 - 15 years; adult = 16 - 86 years \*Adjusted for age, sex, BMI (children model adjusted for BMI percentile and adult model adjusted for absolute BMI), genotype and deprivation

## Discussion

To our knowledge, this is the first national-scale study focusing on the impact of living close to a PCS on lung function, PEx, and fungal and bacterial disease in pwCF. There is limited prior evidence on the health impacts of living near PCSs in the general population or in sub-groups affected by respiratory diseases. Herr *et al* found significant associations between bronchitis (OR = 3.59, 95%CI; 1.40, 9.47), waking up due to cough (OR = 6.59, 95%CI; 2.57,17.7) and coughing during the day (OR = 3.18, 95%CI; 1.24,8.36) among individuals living within 150-200m of a PCS compared to individuals living farther away (>400-500m) in Germany (Herr et al., 2003). In Finland, Aatamila *et al* also found more shortness of breath (OR 1.5, 95% CI 1.0–2.2) and tiredness (1.5, 1.1–2.0) among individuals living within distance zones of <1.5km from waste treatment centres (for municipal waste or composting) compared to individuals living within 3.0 to 5.0 km (Aatamila et al., 2011). Various community and occupational studies have explored the impact of bioaerosols on respiratory health which have been summarised in a systematic review (Pearson et al., 2015a). These studies have shown a possible increased airway irritation among residents living near composting sites. Other studies also found a significant association between organic dust and lung function (Jacobsen et al., 2008; Senthilselvan et al., 2010). Nevertheless, Douglas *et al* (Douglas et al., 2016) did not find an increased risk of respiratory-related hospital admission among the general population living near PCSs in England, using distance from PCS as a proxy for bioaerosol exposure. In a follow-up study, Roca-Barcelo *et al (Roca-Barcelo et al., 2020)* did not find an increased risk of being hospitalised with CF, nor any respiratory-related health outcome, in areas with higher exposure to *A.fumigatus* (estimated using dispersion models) near PCSs in England.

Although we found no associations for children with CF, our study found a relatively lower (1.07%) ppFEV_1_ and higher (1.02 days/year) #IVdays among adults with CF living closer (≤ 4km) to a PCS. To contextualise this, lower FEV1 is strongly associated with higher mortality and poor quality of life, hence it is important to define the percentage of change in FEV_1_ that is clinically relevant and significant in pwCF. FEV1 has been used as the primary outcome in the majority of CF clinical trials due to its reproducibility and reliability (Ramsey et al., 2011; Wainwright et al., 2015). For example, Wainwright *et al* reported a significant improvement in ppFEV_1_ ranging between 2.6-4.0% (*p* <0.001) in phase 3, randomized, double-blind study assessing the effects of lumacaftor/ivacaftor among 1,108 CF patients (Wainwright et al., 2015). PEx are associated with a decline in FEV_1_ and poor quality of life. Furthermore, they are considered a major cause of morbidity among pwCF. Although small reductions in mean ppFEV_1_ and a one day reduction in #IV days may be not clinically significant at an individual level, this could represent an important difference at the population level in the UK CF population. Environmental risk factors may play a critical role in the development of PEx with a significant consequences and serious implications for the health of pwCF. A better understanding of these environmental risk factors, including how they mediate adverse effects and influence disease progression, could provide insight into the pathogenesis of respiratory disease in pwCF.

There was a significant reduction in mean ppFEV_1_ (5.2%) and (4.2%) in CF adults with *A.fumigatus* and *Candida spp* positive cultures living close to a PCS, respectively. Our study cannot tell us the origin of the pathogens, the potential mechanism underlying this, or indeed if it relates to an unmeasured factor. One possible explanation could be a higher sputum fungal burden (density) (Williams et al., 2019a) close to PCSs. Williams *et al* found high anthropogenic *A.fumigatus* modelled concentrations in a 3-4 km radius around some composting facilities in England (Williams et al., 2019a). Although not statistically significant, living close to a PCS was associated with a higher prevalence of ABPA in children, which may provide some indirect evidence in support of this theory, as a higher density of fungal spores in the atmosphere could lead to greater sensitisation. This requires testing in robust prospective studies. Quantitative molecular microbiological methods to identify fungi at species level would be able to elucidate fungal burden in sputum (Cuthbertson et al., 2021). The majority of studies to date have only explored the association between lung function and positive sputum fungal cultures by traditional culture-based methods (Burgel et al., 2016a; Kraemer et al., 2006). For example, Amin *et al* reported mean FEV_1_ values of 79.2% and 86.1% among pwCF with *Aspergillus* positive and negative cultures, respectively (*p=*0.04) (Burgel et al., 2016a), while Kraemer *et al* found a decline in lung function in CF children with positive *Aspergillus* cultures compared to CF controls.

The major strengths of this UK-wide study are the large number of pwCF studied, the high quality of the UK CF Registry data, the availability of detailed individual-level residential information (full residential postcode) to assess distances, the inclusion of all registered PCS in the UK, the use of an improved genotype classification, and the inclusion of multiple covariates. This study aimed to consider proximity to PCS as a proxy for bioaerosol exposure and associations with selected outcomes in pwCF; therefore, we did not account for wind speed/direction, topography, or bioaerosol dispersion. We were also unable to account for background levels of bioaerosol (including seasonal variation), or other potential sources of anthropogenic bioaerosol (e.g. sewage treatments works, intensive farming). Future studies should consider using more detailed exposure assessment methods. There is also potential for further exposure misclassification as we geocoded PCSs based on postcodes provided by Environment Agencies which may not always reflect the exact location of the PCSs. We also did not differentiate between PCS by size or site type (e.g. whether the PCS is open windrow (outdoors) where emissions are uncontained, or in-vessel (indoors) where emissions may be contained), which would likely influence exposure, as data on these characteristics provided by the Environment Agencies were not complete. Bioaerosols emitted from composting facilities are likely to return to background levels within a few kilometres (Williams et al., 2019b). Due to the small number of pwCF living within short distances, we were unable to investigate risk in this zone; nor within 250 m as no pwCF lived within this distance from a PCS. Therefore, we were unable to investigate risks where bioaerosol concentrations are likely to be the highest (Deacon et al., 2009; Douglas et al., 2016; Pankhurst et al., 2011; Pearson et al., 2015b). Household measurements or close monitoring of the few pwCF living close to PCS may provide quantitative evidence to support or adjust the UK Environment Agency’s current risk assessment limit (250m).

As with any registry study, there is the potential for bias, particularly by unmeasured factors. The small decline in ppFEV_1,_ the small increase in #IVdays and the associations found for fungal and bacterial disease in adults living within 4 km of a PCS may be due to residual confounding because the characteristics of those living near a PCS could be different from those living farther away. We limited our analyses to the most common fungal and bacterial species, so other potentially important clinical factors could have been missed. Further analyses should account, for example, for the use of antifungal treatment. Sputum results were based on UK CF Registry criteria and sampling frequency and technique (i.e. sputum, cough swab and bronchoscopy) will have varied (e.g. a higher proportion of children will have provided cough swabs). Further work including longitudinal sampling and molecular microbiological methods would be of benefit as this could help to: i) better characterise fungal infections experienced in pwCF; ii) investigate whether pwCF who have higher exposure to bioaerosols have an increased fungal burden density); and iii) explore the respiratory mycobiome in pwCF, including fungal diversity and its association with seasonal factors.

## Conclusions

We found associations between proxies of disease severity in pwCF living within 4km from a PCS (as a proxy for bioaerosol exposure) compared to those living farther away. The clinical relevance of these findings needs to be further assessed by studies with a better measure of bioaerosol exposure than the distance from the site, which accounts for bioaerosol dispersion, longitudinal sampling and molecular methods to identify and quantify pathogens in sputum. If our results are confirmed, the associations identified could have important implications on: i) the living environments of pwCF, ii) clinical advice to pwCF; iii) public health advice provided to vulnerable populations living near PCS; and iv) how PCS are regulated and permitted.

## Data Availability

This data is not publicly available due to the sensitive clinical information of the study participants confidentiality however anyone who requires it can contact the UK Cystic Fibrosis Registry directly.

## Acknowledgements

We thank Rebecca Cosgriff, Elaine Dunn and the UK CF Registry team for the help obtaining and using CF data, and the patients and clinicians feeding the database. We thank Hima Daby, Gajanan Natu and Eric Johnson of the SAHSU database team for technical support. We are grateful to the UK Environment Agency, Natural Resources Wales in Wales, Scottish Environment Protection Agency in Scotland and Northern Ireland Environment Agency in Northern Ireland for the composting sites data and their technical input. We thank Andrew Jones for his thoughtful guidance on the project.

## Supplementary data

## Appendix A: Allergic Bronchopulmonary Aspergillosis and Candida spp

ABPA is defined using standard international diagnostic criteria, including acute or subacute clinical deterioration (cough, wheeze, exercise intolerance, exercise-induced asthma, change in pulmonary function, or increased sputum production) not attributable to another aetiology, total IgE > 500 IU/ml and a positive skin prick test for *Aspergillus* antigen (> 3 mm) or positive specific IgE for *A.fumigatus*. Also, either precipitating to *A.fumigatus* or *in vitro* demonstration of IgG antibodies to *A.fumigatus* or new or recent abnormalities on chest radiography (infiltrates or mucus plugging) or chest CT (characteristic changes) that have not cleared with antibiotics and standard physiotherapy^1^. The point prevalence of ABPA was 8.8% among PwCF living close to a PCS compared to 7.9% in those living farther away (p= 0.397).

## Appendix B: Distance bands

In line with previous studies^2-4^, we considered multiple distance bands around each PCS (Table S1). Analyses were conducted for each of the following distance bands (≤250m; 250-750m; 750m-1,5km; 1,5-4,0km and >4,0km). Only results for within and beyond 4km are presented due to the lack of statistical power due to small numbers of pwCF living in the smaller radii.

## Appendix C: Rural-urban

In England and Wales, all urban major/ minor conurbations (population of 10,000 or more) and urban cities are classified into urban area^5^. Rural towns, fringes, villages, hamlets and isolated dwellings were classified as rural area. We have extracted rural and urban areas classification from ONS postcodes directly based on residential postcode (https://geoportal.statistics.gov.uk/). In Scotland, rural areas are classified as an area with a settlement of fewer than 3,000 people and settlement of over 3,000 people is classified as urban. In Northern Ireland, the urban area is defined as an area with a population of 5,000 or more and an area with a population of less than 5,000 is classified as rural.

