## supplementary for "Assessing the health risk of living near composting facilities on lung health, fungal and bacterial disease in cystic fibrosis: a UK CF Registry study"

**Supplementary Tables**

**Table S1.** Number of people with CF (pwCF) living within specific distances from a permitted composting site (PCS) in the UK in 2016

| *Distance to a PCS* | *pwCF* | *%* |
| --- | --- | --- |
| 0-250 m | 0 | 0.0 |
| > 250 - ≤ 750 m | 18 | 0.2 |
| > 750 - ≤ 1.5 km | 89 | 1.0 |
| > 1,5 - ≤ 2.5 km | 260 | 2.8 |
| > 2.5 - ≤ 4.0 km | 649 | 6.9 |
| > 4.0 km | 8,345 | 89.1 |
| TOTAL | 9,361 | 100.0 |

**Table S2:** Classification of *CFTR* genotypes with frequencies among pwCF (n=9,361) in the UK in 2016

| SNO | Classification | Description | n | % |
| --- | --- | --- | --- | --- |
| 1 | **Homozygous F508del** | F508del on both alleles. | 4,603 | 49.2 |
| 2 | **F508del/ Minimal function (MF)** | F508del on one allele and any minimal function mutation on the other allele i.e a mutation that produces either no protein or protein that does not respond to currently approved *CFTR* modulators, | 1,822 | 19.5 |
| 3 | **Residual function (RF)/other** | RF mutations include 2789+5G>A, 3272-26A>G, 3849+10kbC>T, 711+3A->G, A1067T, A455E, D110E, D110H, D1152H, D1270N, D579Gx, E193K, E56K, E831X, F1052V, F1074L, G1069R, K1060T, L206W, P67L, R1070Q, R1070W, R117C, R347H, R352Q, R74W, S945L and S977F  RF on one allele and any other mutation on the 2nd allele were counted in this group including RF/RF, F508del/RF, MF/RF, RF/Gating and RF/Other. This group also includes R117H. | 1,070 | 11.4 |
| 4 | **Gating (G)/any*** | G551D, G1244E, G1349D, G178R, G551S, S1251N, S1255P, S549N, or S549R. Gating on one allele and any other mutation on the 2nd allele including F508del/G, G/G, G/Other and MF/G but not RF/G (see above) | 583 | 6.2 |
| 5 | **MF/MF** | All genotypes with MF mutations on both alleles | 298 | 3.2 |
| 6 | **F508del/Other** | F508del on one allele and the second allele was either not identified or identified but classification into the above groups was not possible | 663 | 7.1 |
| 7 | **MF/other** | MF on one allele and the second allele was either not identified or identified but classification into the above groups was not possible | 146 | 1.6 |
| 8 | **Other/other** | On both alleles either the mutation was not identified, or it was identified but classification into the above categories was not possible. | 176 | 1.9 |

*****Excludes Gating/Residual

**Table S3:** Linear regression analysis of the relationship between ppFEV_1_ and residential postcode distance from PCS among children and adults with CF in the UK in 2016. p-value <0.05 are shown in bold

| **ppFEV_1_** | **Children (6 -15 years)**  (n=2,370) | | **Adult (16 - 86 years)**  (n=5,430) | |
| --- | --- | --- | --- | --- |
|  | ≤ 4 km  (n=222) | > 4 km  (n=2148) | ≤ 4 km  (n=644) | > 4 km  (n=4786) |
| **Unadjusted**  β (95%CI) | -0.75 (-2.45, 0.96) | Ref | -1.44 (-2.84, -0.04) | Ref |
| *p value* | 0.391 | | **0.044** | |
| AIC | 19297.9 | | 47438.5 | |
| **Adjusted model I**  β (95%CI) | -0.90 (-2.54, 0.74) | Ref | -1.47 (-2.80, -0.14) | Ref |
| *p value* | 0.282 | | **0.031** | |
| AIC | 19118.11 | | 46929.3 | |
| **Adjusted model II**  β (95%CI) | -1.11 (-2.69, 0.47) | Ref | -1.50 (-2.78, -0.22) | Ref |
| *p value* | 0.168 | | **0.022** | |
| AIC | 18935.0 | | 46535.8 | |
| **Adjusted model III**  β (95%CI) | -1.06 (-2.63, 0.51) | Ref | -1.34 (-2.60, -0.10) | Ref |
| *p value* | 0.184 | | **0.034** | |
| AIC | 18910.4 | | 46185.4 | |
| **Adjusted model IV**  β (95%CI) | -1.00 (-2.56, 0.57) | Ref | -1.20 (-2.42, 0.03) | Ref |
| *p value* | 0.211 | | 0.056 | |
| AIC | 18896.5 | | 46062.6 | |
| **Adjusted model V**  β (95%CI) | -0.71 (-2.8, 0.87) | Ref | -1.05 (-2.28, 0.17) | Ref |
| *p value* | 0.377 | | 0.092 | |
| AIC | 18887.7 | | 46041.3 | |
| **Adjusted model VI**  β (95%CI) | -0.74 (-2.31, 0.83) | Ref | -1.07 (-2.29, 0.16) | Ref |
| *p value* | 0.343 | | 0.088 | |
| AIC | 18887.6 | | 46040.6 | |

ppFEV_1_ = percent predicted FEV_1_; CF = Cystic fibrosis; PCS = permitted composting site

| **Children**  Model I adjusted for BMI percentile  Model II adjusted for BMI percentile and age,  Model III adjusted for BMI percentile, age and mutation  Model IV adjusted for BMI percentile, age, mutation and *P.aeruginosa*  Model V adjusted for BMI percentile, age, mutation and *P.aeruginosa* and deprivation  Model VI adjusted for BMI percentile, age, mutation and *P.aeruginosa,* deprivation and sex | **Adults**  Model I adjusted for *P.aeruginosa*  Model II adjusted for *P.aeruginosa* and absolute BMI  Model III adjusted for *P.aeruginosa* ,absolute BMI and age  Model IV adjusted for *P.aeruginosa* ,absolute BMI, age and mutation  Model V adjusted for *P.aeruginosa* ,absolute BMI, age, mutation and deprivation  Model VI adjusted for *P.aeruginosa* ,absolute BMI, age, mutation, deprivation and sex |
| --- | --- |
